# The Desire to Avoid Pregnancy Scale: clinical considerations and comparison with other questions about pregnancy preferences

**DOI:** 10.1101/2022.11.07.22281888

**Authors:** Jennifer A Hall, Geraldine Barrett, Judith Stephenson, Natalie Edelman, Corinne H Rocca

**Affiliations:** Reproductive Health Research Department, UCL Elizabeth Garrett Anderson Institute for Women’s Health, London, UK; School of Sport & Health Sciences, University of Brighton, Brighton & Hove, UK; Primary Care & Public Health, Brighton & Sussex Medical School, Brighton & Hove, UK; Advancing New Standards in Reproductive Health (ANSIRH), Department of Obstetrics, Gynecology and Reproductive Sciences, University of California, San Francisco (UCSF) School of Medicine, Oakland, CA 94612, USA

**Keywords:** Pregnancy planning, pregnancy prevention, sensitivity, specificity, prediction, desire to avoid pregnancy

## Abstract

**Background:** Clinicians and people of reproductive age would benefit from a reliable way to identify who is, or is not, likely to become pregnant in the next year, to direct health advice. The 14-item Desire to Avoid Pregnancy (DAP) Scale is predictive of pregnancy; this paper compares it with other ways of assessing pregnancy preferences to shortlist options for clinical implementation.

**Methods:** A cohort of 994 UK women of reproductive age completed the DAP and other questions about pregnancy preferences, including the Attitude towards Potential Pregnancy Scale (APPS), at baseline, and reported on pregnancies quarterly for a year. For each question, DAP item, and combinations of DAP items, we examined the predictive ability, sensitivity, specificity, area under the receiver operating curve (AUROC), and positive and negative predictive values.

**Results:** The AUROCs and predictive ability of the APPS and DAP single items were weaker than the full DAP, though all except one had acceptable AUROCs (>0.7). The most predictive individual DAP item was ‘It would be a good thing for me if I became pregnant in the next three months’, where women who strongly agreed had a 66.7% chance of pregnancy within 12months and the AUROC was acceptable (0.77).

**Conclusion:** We recommend exploring the acceptability to women and healthcare professionals of asking a single DAP item (‘I wouldn’t mind if I became pregnant in the next three months’), possibly in combination with additional DAP items. This will help to guide the provision of information and services to support reproductive preferences.

**Key messages:**

- Clinicians do not currently have a valid and reliable way of asking people about their pregnancy preferences.
- A single item from the Desire to Avoid Pregnancy Scale is effective at identifying who is likely to become pregnant in the next year; other questions, including the APPS, are less discriminative but may be more acceptable.
- The acceptability to women and health professionals of different ways of asking people about their pregnancy preferences in health care settings should be explored.

## INTRODUCTION

While there are a multitude of measures and screening tools available to predict pregnancy-related conditions, such as pre-eclampsia^1^ or gestational diabetes,^2^ there are no valid and reliable clinical tools to identify women^a^ who are likely to become pregnant.^1^ Such a tool would have great utility for clinicians working with women of reproductive age as it could be used to guide discussions on either preparation for pregnancy or contraceptive options, depending on if and when a pregnancy might be desired.

The Desire to Avoid Pregnancy (DAP) Scale was developed in the United States (USA) in 2019,^3^ and validated in the United Kingdom (UK) in 2022, where it was also shown to be highly predictive of pregnancy, with women with the lowest DAP score having an 80% chance of becoming pregnant within 12 months.^4^ Subsequently the sensitivity and specificity of the DAP have been described (0.78 and 0.81 respectively, at a cut-point<2)^5^ and further analysis has shown how DAP score is associated with socio-demographic factors,^5^ findings which are in keeping with the literature on pregnancy preferences.^6-8^

While there may be ways of incorporating the DAP scale into clinical practice, e.g., pre-consultation digital use or self-completion, a tool with 14-items is likely to be impractically lengthy in many clinical consultations. Use of selected items from the DAP or other questions, may provide a brief and viable alternative if they distinguish sufficiently between who will and who will not become pregnant. While other, shorter ways of assessing pregnancy preferences exist, such as the Attitude toward Potential Pregnancy Scale (APPS)^9^ and the ‘One Key Question^®^’ (OKQ),^10^ there are no data on their predictive ability and, until the DAP scale, there had not been a ‘gold standard’ to compare them to.

The aims of this paper are to: 1) evaluate the predictive ability, sensitivity, specificity, area under the receiver operating curve (AUROC) and positive and negative predictive values (PPV and NPV) of different methods of assessing pregnancy preferences; and 2) examine the same criteria for individual DAP items and selected combinations of DAP items. Using the findings of these two aims, options are shortlisted for consideration for clinical implementation.

## METHODS

### Sample

We analysed data from a cohort of 994 non-pregnant women, aged 15 and over, who were recruited in the UK in October 2018 and followed up every three months for one year. The full details of recruitment and participation are described elsewhere^4^ but, in brief, people who self-reported as female, were pre-menopausal and not sterilised were recruited though advertising at a school, a university, a sexual health clinic and a pregnancy termination clinic, as well as through online recruitment via both paid advertisements (Instagram and Facebook) and sharing through networks.^4^ The survey was programmed in RedCap^11,12^ and included the DAP scale, other questions about pregnancy preferences and socio-demographics. At each quarterly follow-up, participants were asked whether they were currently pregnant or had been pregnant since the last survey. The women who did not complete 12month follow-up were not significantly different in sociodemographic characteristics from those who did (suggesting that there is no selection bias in the loss to follow up).^5^

### Measures

#### Outcome

We created a binary variable of any incident pregnancy between baseline and 12months. Given low attrition and to ease interpretation, we included participants in pregnancy denominators until they were lost to follow-up and report percentages rather than rates.

#### Attitude toward Potential Pregnancy Scale

The Attitude toward Potential Pregnancy Scale (APPS) is a five-item measure of a woman’s emotional outlook regarding a potential pregnancy (Table 1).^9^ Each item is scored one-to-five on a visual anchored scale and summed to give an overall score from five to 25; higher scores represent a more positive attitude to pregnancy. The APPS had a Cronbach’s alpha of 0.86 among 130 participants in the USA but has not been examined in the UK.

**Table 1.**
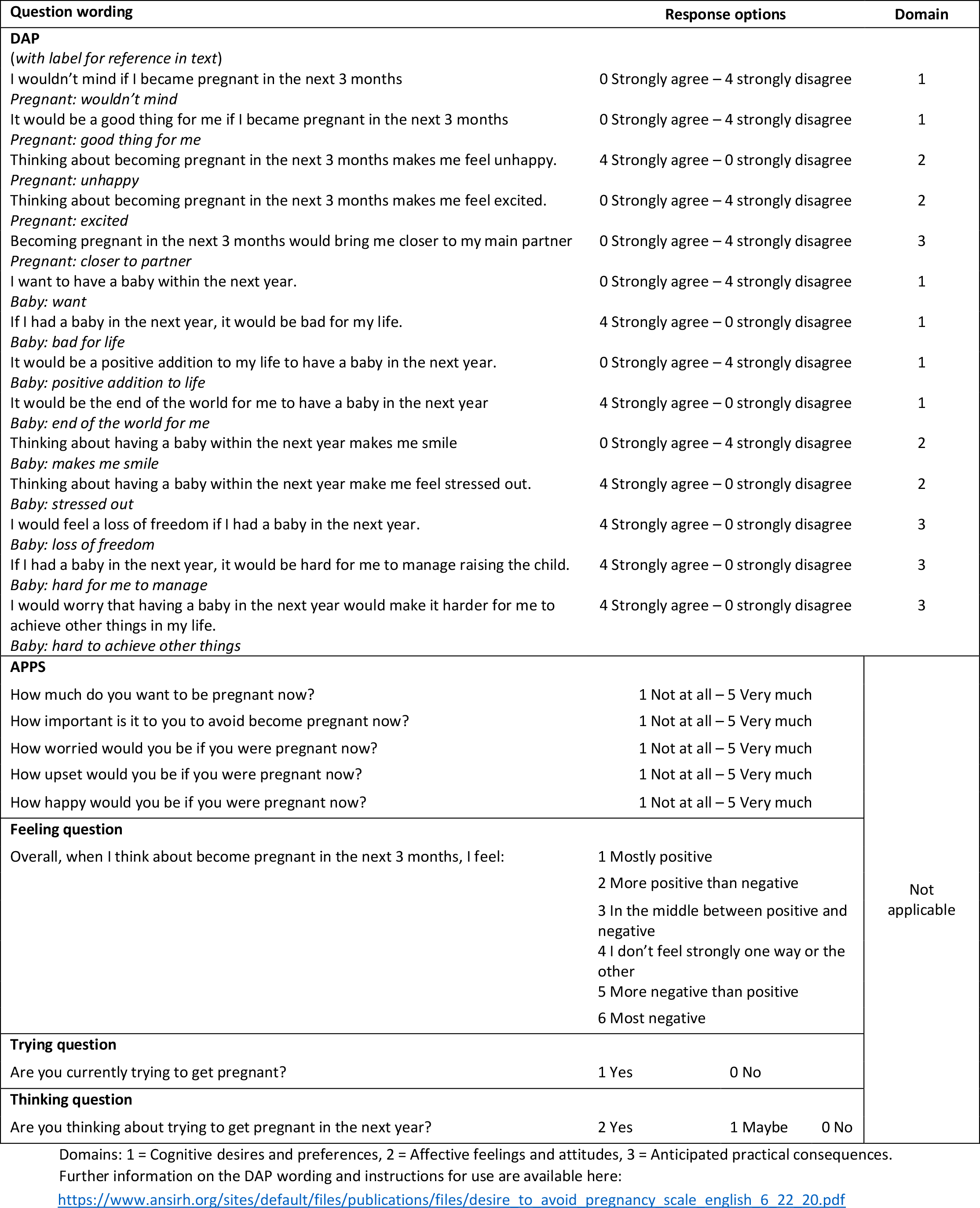
Wording of questions about preferences regarding future pregnancy.

#### Other pregnancy preference questions

We compared the DAP score with three other questions asking about pregnancy preferences (Table 1). The ‘feelings question’ was used in the ADAPT study;^13^ the ‘trying’ and ‘thinking’ questions are ones that clinicians in the UK have told us are the kind of questions they currently use. We did not use the OKQ^®^ ‘Would you like to become pregnant in the next year?’ as this is a proprietary tool that is unlikely to be available for widespread implementation in the UK, but the ‘thinking’ question is similar.

#### DAP Scale

The DAP scale is a psychometrically validated measure covering three conceptual domains: 1) cognitive desires and preferences; 2) affective feelings and attitudes; and 3) anticipated practical consequences (Table 1).^3^ Each item uses a Likert scale, scored zero-to-four, to ask women how much they agree or disagree with a statement about either becoming pregnant in the next three months or having a baby in the next year. The DAP was developed following an extensive item development process and item response theory was utilised to create the final tool. Responses are averaged, producing a total score between zero and four; higher scores represent a higher desire to avoid pregnancy.

### Plan of Analysis

#### Aim 1: Evaluating the performance of other methods of assessing pregnancy preferences

We assessed the APPS’s reliability (internal consistency) with Cronbach’s alpha (>0.7 considered acceptable)^14^ and checked that all item-rest correlations were positive and >0.2.^15^ We examined construct (structural) validity via principal components analysis and considered the scale structurally valid if all items loaded on to one component with an Eigenvalue >1.^16^

To compare the assessment of pregnancy preferences by the DAP versus the APPS and the three individual questions (‘feeling’, ‘trying’ and ‘thinking’), we examined the relationships among them. As the DAP and the APPS yield continuous scores, we used the Pearson correlation coefficient to examine the strength of association, expecting a negative correlation (i.e., as DAP score increases, APPS score decreases), considering a strong correlation to be lower than -0.7. This also served as a test of the APPS’s concurrent validity. The range of DAP scores within, and distribution of DAP scores across, the response options of each question was examined. The Kruskall-Wallis test was used to assess whether differences in median DAP score across response options within each question were significant.

To investigate how well each approach predicted actual pregnancy, we modelled the probability of pregnancy using logistic regression models and examined the sensitivity, specificity, AUROC, PPV and NPV, using the Youden index suggested cut-point.^17^ The AUROC represents the tool’s ability to identify who will become pregnant. An AUROC of 0.7-0.8 was considered acceptable, 0.8-0.9 excellent and >0.9 outstanding.^18^

#### Aim 2: Examination of the performance of individual and combinations of DAP items

We explored the predictive ability of each individual DAP item, as well as selected combinations of items, using logistic regression, with incident pregnancy as the outcome. The item combinations were designed to ensure coverage of the three conceptual domains, positively and negatively framed items, different time frames and use of ‘pregnancy’ and ‘baby’, as well as the predictive ability of the items. The combinations were cross-validated by developing them on one half of the data (created with a random split) and tested on the other half to avoid over-fitting and give a more accurate estimate of how the questions perform outside the data they were developed from.^19 20^ The results from the testing data are presented. The predictive ability, sensitivity, specificity, AUROC, PPV and NPV of the selected items/combinations were calculated (using the Youden index suggested cut-point)^17^ and compared with the total DAP scale.

To inform which question(s) provided the best balance between brevity and predictive ability, the performance of the single questions, the APPS, and selected DAP items/combinations were compared using the predictive ability, AUROC and number of items/questions, to make recommendations on which item/question(s) should be taken forward for further consideration.

### Patient and Public Involvement

We undertook public involvement in the development of our overall programme of research on pregnancy planning; the P3 Study. Findings were frequently discussed with the P3 Study’s PPI group to inform the next steps and the group is currently planning our wider dissemination.

## RESULTS

### Sample

As previously described,^4^ the baseline cohort of 994 women were aged 15-50 years (median=31, IQR=23-36, mean=29.7). Most (82%) were in a relationship, 82% described themselves as heterosexual and 84% identified as white. The sample was quite highly educated with 39% having an undergraduate degree and 31% a postgraduate or professional qualification. 57% had one or more children in their household. The dataset is available in the UCL Research Data Repository.^21^

#### Aim 1: Evaluating the performance of other methods of assessing pregnancy preferences

The full range of APPS scores (5-25) was reported in our cohort. The Cronbach’s alpha for the APPS was 0.93, all item-rest correlations were >0.2 and positive, and all items loaded on to one component with an Eigenvalue of 3.87. The Pearson correlation coefficient between the APPS and DAP was -0.893 (p<0.001) showing a strong negative correlation (i.e., as a woman’s attitude towards pregnancy becomes more positive her desire to avoid pregnancy reduces.) The median DAP score was statistically significantly different across the response options for the APPS and each of the ‘feeling’, ‘trying’ and ‘thinking’ questions (p-values in Table 2). For example, women who felt ‘mostly positive’ about pregnancy had a median DAP score of 0.64 (low desire to avoid pregnancy) whereas women who felt ‘mostly negative’ about pregnancy had a median DAP score of 3.50 (high desire to avoid pregnancy).

**Table 2.**
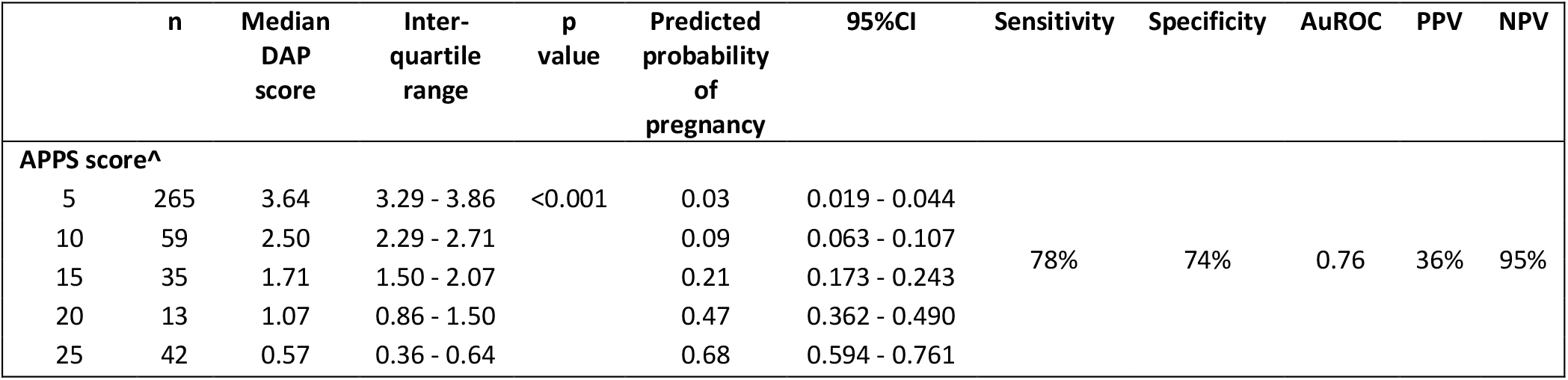
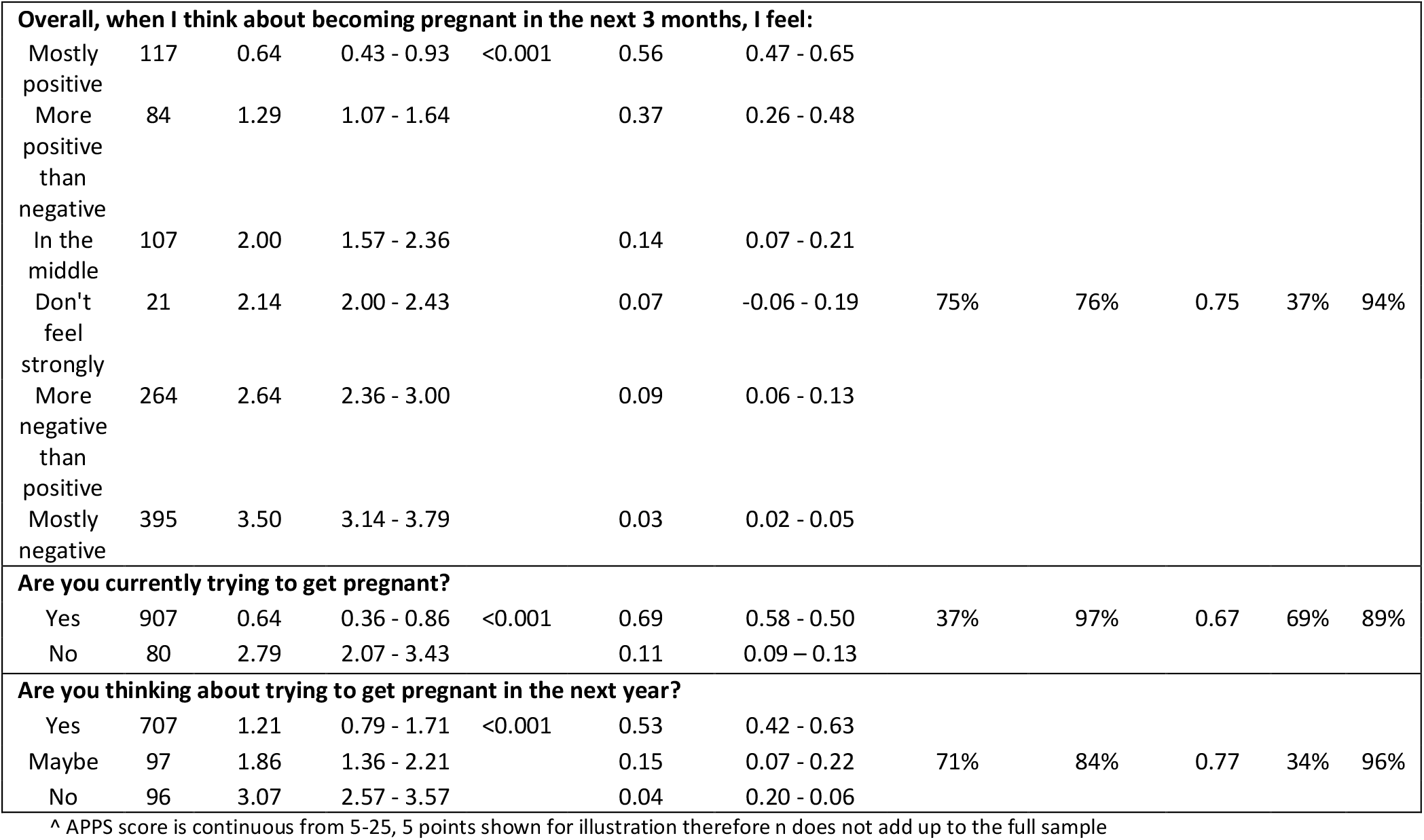
Relationship with DAP and predicted probability, sensitivity, specificity, AuROC, PPV and NPV of other questions.

The AUROCs and predictive ability of the APPS and the single questions were weaker than the full DAP (AUROC 0.87, predictive ability 79.4%), though all except the ‘trying’ question had acceptable AUROCs of >0.7.

#### Aim 2: Evaluating the performance of individual and combinations of DAP items

All DAP items and domains were associated with pregnancy. The best performing individual DAP item in terms of pregnancy prediction was ‘Pregnancy: good thing for me’, where there was a 66.7% chance of pregnancy within 12months among women who strongly agreed (Table 3). At a cut point of 2.5 this item also had an acceptable AUROC (0.77).

**Table 3.** Exploration of the predicted probability, sensitivity, specificity, AuROC, PPV and NPV of selected individual and combined DAP items.

| Description | Predicted probability of pregnancy at DAP score | Empirical cut-point | Sensitivity | Specificity | AuROC | PPV | NPV | Domains covered | Positive & Negatively worded items |
| --- | --- | --- | --- | --- | --- | --- | --- | --- | --- |

|  | 0 | 4 |  |  |  |  |  |  |  |  |
| --- | --- | --- | --- | --- | --- | --- | --- | --- | --- | --- |
| Total DAP:<br>*All 14 items* | 79.4% | 0.9% | 1.96 | 78.0% | 81.0% | 0.85 | 43.0% | 95.0% | All | Yes |
| Single DAP item:<br>*Pregnant: good thing for me.* | 66.7% | 2.1% | 2.5 | 68.1% | 86.3% | 0.77 | 49.0% | 93.3% | 1 | Positive only |
| Single DAP item:<br>*Baby: end of the world for me.* | 27.4% | – | 0.5 | 65.3% | 77.7% | 0.72 | 36.2% | 92.0% | 1 | Negative only |
| Two DAP items:<br>*Pregnant: good thing for me & Baby: end of the world for me.* | 71% | 0.7% | 1.25 | 65.3% | 90.1% | 0.78 | 56.0% | 93.1% | 1 | Yes |
| Three DAP items: *Pregnant: good thing for me, Pregnant: excited, Baby: hard to achieve other things.* | 83% | 2.8% | 2.15 | 62.5% | 87.1% | 0.75 | 48.4% | 92.3% | All | Yes |
| Three DAP items: *Pregnant: good thing for me, Pregnant: excited, Baby: end of the world for me.* | 73% | 1.2% | 1.49 | 66.7% | 87.4% | 0.77 | 50.5% | 93.1% | 1,2 | Yes |
| Three DAP items: *Pregnant: good thing for me, Baby: end of the world for me, Baby: makes me smile.* | 68% | 1.0% | 1.49 | 69.4% | 84.4% | 0.77 | 46.3% | 93.5% | 1,2 | Yes |
| Three DAP items: *Pregnant: good thing for me, Baby: want, Baby: end of the world for me.* | 72% | 13.9% | 1.49 | 66.7% | 90.3% | 0.79 | 57.1% | 93.3% | 1 | Yes |
| Three DAP items: *Pregnant: good thing for me, Baby: end of the world for me, Baby: hard for me to manage.* | 76% | 8.8% | 1.82 | 75.0% | 77.2% | 0.76 | 38.8% | 94.1% | 1,3 | Yes |
| Four DAP items:<br>*Pregnant: good thing for me, Pregnant: excited, Baby: want, Baby: end of the world for me.* | 73% | 1.4% | 1.63 | 66.7% | 89.2% | 0.78 | 54.5% | 93.2% | 1,2 | Yes |
| Four DAP items:<br>*Pregnant: good thing for me, Pregnant: excited, Baby: end of the world for me, Baby: hard to achieve other things.* | 81% | 1.2% | 1.88 | 66.7% | 83.9% | 0.75 | 44.4% | 92.9% | All | Yes |
| Six DAP items:<br>*Pregnant: wouldn't mind, Pregnant: unhappy, Baby: want, Baby: positive addition to life, Baby: makes me smile, Baby: hard for me to manage.* | 73% | 1.6% | 1.93 | 75.0% | 80.9% | 0.78 | 43.2% | 94.4% | All | Yes |

Adding a second item of ‘Baby: end of the world for me’ improved the specificity and PPV, without affecting the AUROC. The highest AUROC (0.70) was achieved with the combination of three items ‘Pregnancy: good thing for me’, ‘Baby: want’ and ‘Baby: end of the world for me’, which also had the highest PPV (57.1%). The item combinations had AUROCs between 0.77-0.79, in line with that of the individual item ‘Pregnancy: good thing for me’ suggesting little additional gain.

## DISCUSSION

This is the first examination of the predictive ability of two measures and three single questions about pregnancy preferences on a sample that is broadly representative of women of reproductive age in the UK. Good correlation was previously seen between DAP score and ‘One Key Question^®^’ (OKQ) in the USA.^22^ As that study noted, DAP scores ranged widely within each OKQ response option; women who responded to the OKQ with ‘want to get pregnant in future but not in next year’ had DAP scores ranging from 1–4. Our data demonstrated the same patterns; women who reported feeling ‘mostly negative’ towards pregnancy had DAP scores ranging from 0.286–4, this shows the nuance that the DAP can capture and demonstrates the heterogeneity missed by a single question.

In terms of the AUROC and the predicted probability of pregnancy in the next year, the performance of the APPS and the other questions was weaker than the complete DAP, though to varying degrees. The APPS and the other questions are all shorter than the DAP and therefore potentially less burdensome for clinical use. Arguably, their poorer performance is offset by their brevity. Our data show that the single question ‘Are you currently trying to get pregnant?’ had low sensitivity (37%) and the lowest AUROC (0.67). While women answering yes to this question were highly likely to become pregnant (PPV 69%) there were more pregnancies among women who answered no. Questions like this, which force women to answer yes-or-no, fail to recognise the complexity of the concept of pregnancy preferences.^23^

When considering which question(s) might be best for clinical use, the trade-off between sensitivity and specificity, and the PPV and NPV is important. In addition, the number of questions that need to be asked, and the complexity of combining them to achieve a score, could be a barrier in consultations, therefore having fewer questions will likely increase uptake. As our analysis shows, single questions generally have lower AUROCs than a set of questions, however selected items from the DAP, which have been developed based on rigorous theoretical groundwork, do have higher AUROCs than the less carefully constructed questions. Neither the DAP nor the APPS were designed to be spoken, whereas the other questions lend themselves more easily to being asked verbally.

### Strengths and limitations

We used a large, broadly representative dataset^4^ and a validated measure (the DAP) to assess the performance of a range of questions for assessing pregnancy preferences and have provided preliminary evidence that the APPS is valid in the UK. Given the limitations on the OKQ we were not able to include it in our research. While we used a split dataset to conduct cross-validation within our sample, which is a strength, confirmation of our findings in other populations would provide further confidence in selecting the most appropriate question(s). Finally, the selected DAP items/combinations were based on their potential as screening tools for pregnancy, as well as theoretical considerations; item-response theory-based analysis of the psychometric properties of these combinations could be conducted.

## CONCLUSION

Discussions about pregnancy preferences are important, regardless of whether the patient wants to become pregnant in the future or not. Ensuring that those who do not wish to become pregnant have the right support to avoid pregnancy is just as important as identifying those who might benefit from pre-pregnancy health advice or those who may benefit from both. Equally, for those who have never formally considered their preferences, it provides an opportunity to empower them and increase their agency by highlighting that people do have choices about pregnancy and parenthood (recognising the effect of external factors) and encouraging them to explore their aspirations.

In the context of a face-to-face clinical encounter, where the full DAP is less likely to be suitable, a tool to assess people’s preferences regarding a future pregnancy needs to be both practical (short) and discriminative i.e. identify who is and is not likely to become pregnant in the short term so that the appropriate advice can be given. Based on our findings we recommend exploring the acceptability to women and health care professionals of a single item from the DAP (‘I wouldn’t mind if I became pregnant in the next three months’) or a combination of this with two additional DAP items (either ‘Baby: want’ and ‘Baby: end of the world for me’ or ‘Pregnant: excited’ and ‘Baby: hard to achieve other things’) adapted from the written format to a spoken one.

## Supporting information

STROBE Cohort Checklist

## Data Availability

The dataset is available online at the UCL Research Data Repository, doi 10.5522/04/21263007.

## Acknowledgements

We would like to thank all the women who took part in the P3 study, the P3 Study steering committee and Catherine Stewart for her support in preparing the manuscript for submission.

## Contributors

JH: guarantor, conceptualisation, funding acquisition, methodology, investigation, data curation, formal analysis, writing-original draft, visualisation, writing-review and editing, project administration. GB: conceptualisation, methodology, writing-review and editing. JS: conceptualisation, writing-review and editing CR: conceptualisation, methodology, writing-review and editing. NE: methodology, writing-review and editing.

## Data availability statement

The dataset is available in the UCL Research Data Repository, doi 10.5522/04/21263007.

## Footnotes

a Where we refer to ‘women’ this should be taken to include people who do not identify as women but who have the capability to become pregnant.

